# A Genetics-First Approach to Dissecting the Heterogeneity of Autism: Phenotypic Comparison of Autism Risk Copy Number Variants

**DOI:** 10.1101/2020.01.14.20017426

**Authors:** Samuel J.R.A. Chawner, Joanne L. Doherty, Richard Anney, Kevin M. Antshel, Carrie E. Bearden, Raphael Bernier, Wendy K. Chung, Caitlin C. Clements, Sarah R. Curran, Goran Cuturilo, Ania M. Fiksinski, Louise Gallagher, Robin P. Goin-Kochel, Leila Kushan, Raquel E. Gur, Ellen Hanson, Sebastien Jacquemont, Wendy R. Kates, Anne M. Maillard, Donna M. McDonald-McGinn, Marina Mihaljevic, Judith S Miller, Hayley Moss, Milica Pejovic-Milovancevic, Robert T. Schultz, LeeAnne Green-Snyder, Jacob A. Vorstman, Tara L. Wenger, IMAGINE-ID Consortium, Jeremy Hall, Michael J. Owen, Marianne B.M. van den Bree

**Author notes:** See IMAGINE-ID consortium member list.

## Abstract

**Objective:** Certain copy number variants (CNVs) greatly increase risk of autism. We conducted a genetics-first study to investigate whether heterogeneity in the clinical presentation of autism is underpinned by specific genotype-phenotype relationships.

**Methods:** This international study included 547 individuals (12.3 years (SD=4.2), 54% male) who were ascertained on the basis of having a genetic diagnosis of a rare CNV associated with high risk of autism (82 16p11.2 deletion carriers, 50 16p11.2 duplication carriers, 370 22q11.2 deletion carriers and 45 22q11.2 duplication carriers), as well as 2027 individuals (9.1 years (SD=4.9), 86% male) with autism of heterogeneous aetiology. The Autism Diagnostic Interview-Revised (ADI-R) and IQ testing were conducted.

**Results:** The four genetic variant groups differed in autism severity, autism subdomain profile as well as IQ profile. However, we found substantial variability in phenotypic outcome within individual genetic variant groups (74% to 97% of the variance depending on the trait), whereas variability between groups was low (1% to 21% depending on trait). We compared CNV carriers who met autism criteria, to individuals with heterogeneous autism, and a range of profile differences were identified. Using clinical cut-offs, we found that 54% of individuals with one of the 4 CNVs who did not meet full autism diagnostic criteria nonetheless had elevated levels of autistic traits.

**Conclusion:** Many CNV carriers do not meet full diagnostic criteria for autism, but nevertheless meet clinical cut-offs for autistic traits. Although we find profile differences between variants, there is considerable variability in clinical symptoms within the same variant.

## Introduction

Autism is a behaviourally defined condition characterized by deficits in social interaction and communication, as well as the presence of restricted, repetitive behaviours and interests^1^. There is considerable heterogeneity in the clinical presentation of autism, in terms of symptom profile, cognitive function and developmental trajectories^2-5^. Studies of large genotyped cohorts of individuals with autism and typically developing controls have identified several chromosomal copy number variants (CNVs) (deletions and duplications >1 kilobase (kb)^6^) as genetic risk factors for autism^7-12^, and have been demonstrated in clinical settings to have predictive value^13^. Although individually rare, collectively pathogenic CNVs are identified in 15% of patients with neurodevelopmental disability^14^. A number of researchers have advocated that the time is ripe for a reverse strategy based on a genetics-first rather than a phenotype-first approach, in order to better understand the clinical heterogeneity of autism^15-17^.

Deletions and duplications at the 16p11.2 (600kb, break points 4 and 5 (BP4-BP5) critical region 29.6-30.2 Mb, build hg19) and 22q11.2 (3 Mb, break points A and D, critical region 19.0-21.5 Mb, build hg19) loci have been identified as risk factors for autism, both from phenotype-first studies that find these variants occur with greater frequency in cohorts of individuals with autism versus controls^7-9^, and genetics-first studies which find that patients diagnosed with 16p11.2 and 22q11.2 CNVs in medical genetics clinics have an elevated frequency of autism diagnosis^18-25^ relative to the frequency in the general population of 1% ^26,27^. It is important to determine whether these variants confer risk for the same autism phenotype or whether the presentation differs by genotype. The former would indicate that genomic risk for autism has common phenotypic effects, whilst the latter would suggest that genetic heterogeneity underpins clinical heterogeneity. Within the autism field, there is a strong notion that the condition is dissociable by genetics^28,29^, with some researchers using the term “autisms”^30^. Early evidence indicates that the 22q11.2 deletion and duplication may have unique autism profiles^24,31^, however, the profiles of the two groups have not been directly compared within the same study, and hence the differences reported could be due to methodological inconsistencies. For the 16p11.2 locus, it has been reported that duplication carriers with autism have lower IQ compared to deletion carriers with autism^25^, however the autism profiles of the two groups have not been compared. It is also important to investigate the extent to which the autism profile of these variants differs from individuals without these variants who have autism (referred to as heterogeneous autism from here onwards).

Comprehensive clinical phenotyping of individuals with autism-risk genetic variants requires large integrated networks of researchers and clinicians using the same clinical instruments. The present study brings together patient data from several international genetics-first consortia of individuals with rare chromosomal conditions associated with high risk of autism. Individuals with deletions and duplications that span critical regions at the 22q11.2 and 16p11.2 loci were ascertained clinically via medical genetics clinics and patient organisations. We aimed to: 1) Characterise and contrast the phenotypes of different autism risk genetic variants, in terms of autism prevalence, severity, symptom domain profile, subdomain profile and IQ; 2) Investigate whether CNV carriers with autism differ in phenotype from individuals with autism of heterogeneous origins.

## Methods

### Participants

#### Genetics-first cohorts

We identified several clinical research sites and consortia which had independently established genetic-first cohorts, and had utilised the Autism Diagnostic Interview – Revised (ADI-R) ^32^ to assess autism, thus allowing data to be easily combined. Data on 566 clinically ascertained CNV carriers were available but 19 cases were removed due to insufficient genotypic information (n=18) and cohort overlap (n=1). This resulted in 547 CNV carriers (12.3 years (SD=4.2), 54% male); 82 with 16p11.2 deletion, 50 with 16p11.2 duplication, 370 with 22q11.2 deletion and 45 with 22q11.2 duplication was provided from the ECHO (ExperienCes of people witH cOpy number variants https://www.cardiff.ac.uk/mrc-centre-neuropsychiatric-genetics-genomics/research/themes/developmental-psychiatry/echo-study-cnv-research) study, the IMAGINE-ID (Intellectual disability and Mental health: Assessing Genomic Impact on Neurodevelopment http://imagine-id.org/) study, the Hospital neurodevelopmental CNV cohort at the Belgrade University Children’s Hospital Belgrade, the International 22q11.2DS Brain and Behaviour Consortium (http://22q11-ibbc.org/), the Center for Autism Research at Children’s Hospital of Philadelphia, and the 16p11.2 European consortium (http://www.minds-genes.org/Site_EN/index.html) and Simons Variation in Individuals Project (VIP) Consortium (https://simonsvipconnect.org/) (Supplementary Table 1 has full details). Cohort demographics by continent are provided in Table 1. The characteristics of these studies have been described elsewhere^23-25,33-37^.

**Table 1:** Cohort demographics. EU, Europe; US, United States.

| Cohort | Sample size | Sex |  | Age |  | Autism |  |
| --- | --- | --- | --- | --- | --- | --- | --- |
|  |  | n (male) | % (male) | mean | SD | n | % |
| **Genetic first cohorts** |  |  |  |  |  |  |  |
| **16p11.2 deletion** | 82 | 43 | 52% | 9.6 | 3.7 | 35 | 43% |
| EU | 12 | 7 | 58% | 10.3 | 4.4 | 7 | 58% |
| US | 70 | 36 | 51% | 9.5 | 3.6 | 28 | 40% |
| **16p11.2 duplication** | 50 | 35 | 70% | 10.8 | 6.7 | 29 | 58% |
| EU | 17 | 16 | 94% | 12.6 | 8.6 | 13 | 76% |
| US | 33 | 19 | 58% | 9.9 | 5.4 | 16 | 48% |
| **22q11.2 deletion** | 370 | 182 | 49% | 13.4 | 3.4 | 85 | 23% |
| EU | 215 | 102 | 47% | 13.7 | 2.4 | 50 | 23% |
| US | 155 | 80 | 52% | 12.9 | 4.4 | 35 | 23% |
| **22q11.2 duplication** | 45 | 35 | 78% | 10.1 | 4.3 | 20 | 44% |
| EU | 11 | 9 | 82% | 10.3 | 2.8 | 6 | 55% |
| US | 34 | 26 | 76% | 10.0 | 4.7 | 14 | 41% |
| **Heterogeneous autism** | 2027 | 1753 | 86% | 9.1 | 4.9 | 2027 | 100% |
| EU | 848 | 731 | 86% | 8.6 | 4.8 | 848 | 100% |
| US | 1179 | 1022 | 87% | 9.5 | 5.0 | 1179 | 100% |

Carrier status for CNVs at the 16p11.2 (critical region 29.6-30.2 Mb, had to span breakpoints 4 to 5, build hg19) or 22q11.2 (critical region 19.0-21.5 Mb, had to span at least low copy repeat regions A-B as pathogenicity of atypical variants outside the A-B region is uncertain, build hg19) loci was confirmed for all individuals through clinical chromosome microarrays, medical records and/or confirmation in a research laboratory (Full genotype information in Supplementary Table 2). Analysis included individuals ≥4 years old. The study was approved by the appropriate local ethics committees and institutional review boards. Each participant and his or her caregiver, where appropriate, provided informed written consent/assent to participate prior to recruitment.

#### Heterogeneous autism cohort

Data on 2053 individuals with autism, ≥4 years old, was accessed from the Autism Genome Project (AGP) ^38^. These individuals were ascertained via autism diagnostic clinics. Of these 2053 individuals, 26 had CNVs at the 16p11.2 and 22q11.2 loci; 7 with 16p11.2 deletion, 4 with 16p11.2 duplication, 4 with 22q11.2 deletion and 11 with 22q11.2 duplication. Given the small sample sizes we did not compare these groups to the remainder of the AGP cohorts; also, previous work has reported on the phenotype of CNV carriers in the AGP cohort^39^. These individuals were not included in the genomic condition groups given the different ascertainment strategies. The remaining 2027 individuals represent a group of individuals with autism for whom the underlying aetiology is heterogeneous (See Table 1 for demographics). Following previous authors^16^, we refer to this cohort as “heterogeneous autism”, rather than “idiopathic.”

#### Autism assessment

All individuals were assessed using the Autism Diagnostic Interview - Revised (ADI-R)^32^ by a research reliable assessor (further information on assessors and assessment sites in Supplementary Table 3). The ADI-R is a semi-structured interview conducted with the primary caregiver about a child’s symptoms both currently and during early development. Total ADI-R score was used as an index of autism severity^37^. Autism domain scores for “Social Interaction”, “Communication” and “Restricted, Repetitive, and Stereotyped Behaviours (RRBs)” were extracted, as well as autism subdomain scores (further details on ADI-R scores in Supplementary Materials). To meet autism criteria on the ADI-R an individual had to meet the clinical cut-offs on each domain (score of 10 for social, 8 (7 if nonverbal) for communication, and 3 for RRB) and there must also have been evidence of developmental abnormality before the age of 36 months.

#### Cognitive assessment

Full Scale IQ (FSIQ), Verbal IQ (VIQ) and Performance IQ (PIQ) scores were derived from age and developmentally appropriate standardized IQ measures as described elsewhere ^23,25,33,36,40^.

### Statistical Analysis

#### Aim 1

Characterising and contrasting the phenotypes of different autism risk genetic variants

##### Autism prevalence within genetic variant groups

Autism prevalence was determined on the basis of the ADI-R diagnostic algorithm^41^. A logit mixed model was performed to determine whether genetic variant group (22q11.2 deletion, 22q11.2 duplication, 16p11.2 deletion, 16p11.2 duplication) was a predictor of autism diagnosis, whilst accounting for gender and age. Following previous international studies of the 16p11.2 duplication, we included site (European vs United States) as a covariate^25,34^. Post-hoc contrasts were conducted to establish autism prevalence differences between genetic variant groups with Tukey adjustment for multiple comparisons. The percentage of individuals who did not meet autism criteria but did meet the clinical cut-off in one or more domains was additionally calculated.

##### Autism profiles between genetic variant groups

To investigate possible differences in autism profiles between genetic variant groups, a series of analysis of covariance (ANCOVA) models were conducted with group as a predictor and the following phenotypic variables as outcome measures: ADI-R total as an index of *autism severity* (ADI-R total score), *autism domain profile, autism subdomain profile* and *IQ profile*, whilst accounting for gender, age and site (see Supplementary Materials for full information). Tukey’s method was used to conduct post-hoc contrasts between genetic variant groups, producing p-values adjusted for the number of contrasts. Eta-squared values were calculated to estimate the proportion of variance explained by genetic variant group (between group differences). We also calculated the variance that is explained by variable expressivity within the four genetic variant groups, i.e. variance not explained by genetic variant group, age, gender and site. Analyses of ADI-R total score, domain and subdomain scores were repeated including FSIQ as a covariate, to investigate whether differences in autism phenotype were driven by FSIQ.

#### Aim 2

Symptom profiles of individuals with autism within the genetic variant groups and individuals with “heterogeneous autism”

To compare autism in the genetic variant groups to *heterogeneous autism* (i.e., individuals from the AGP dataset who did not have 16p11.2 and 22q11.2 CNVs; n=2027), we conducted analyses leaving out individuals within the genetic variant groups who did not meet ADI-R criteria for autism, and compared the profiles to individuals with heterogenous autism. This resulted in 5 groups: *16p11*.*2 deletion + autism; 16p11*.*2 duplication + autism*; *22q11*.*2 deletion + autism; 22q11*.*2 duplication + autism*; and *heterogeneous autism* (Table 1).

MANCOVA analysis was conducted with group as a predictor and phenotypic scores as the outcomes, whilst accounting for gender, age and site. As in aim 1, analyses were run for *autism severity* (ADI-R total score), *autism domain profile, autism subdomain profile* and *IQ profile*. Post hoc contrasts to investigate the difference between CNV + autism groups in relation to the autism group were conducted with Tukey adjustment for multiple comparisons.

To investigate whether male-to-female ratios differed between the five groups, we used a logit model with gender as a binary outcome, and group as a predictor, whilst taking account of fixed effects of age, and the random effect of site.

For aims 1 and 2 a Benjamini-Hochberg False Discovery Rate (B-H FDR) multiple testing correction of 0.05 was applied to p-values.

## Results

### Aim 1

Characterising and contrasting the phenotypes of different autism risk genetic variants

#### Autism prevalence within genetic variant groups

Within our cohort of CNV carriers ascertained clinically via medical genetics clinics and patient organisations; 43% of individuals with 16p11.2 deletion, 58% of individuals with 16p11.2 duplication, 23% of individuals with 22q11.2 deletion and 44% of individuals with 22q11.2 duplication met ADI-R criteria for autism (Table 1). Genetic variant group was a significant predictor of autism diagnosis (p<0.001). Post-hoc contrasts revealed that autism prevalence in the 22q11.2 deletion carrier group (23%) was significantly lower compared to the 16p11.2 deletion (43%, p=0.004), 16p11.2 duplication (58%, p<0.001) groups; the remaining genetic variant group differences were not significant.

Within CNV carriers who did not meet formal autism diagnosis, we examined the proportion who met clinical cut-off criteria for one or more domains on the ADI-R. Amongst the 378/547 (69%) individuals who did not meet criteria for autism, 205/378 (54%) were found to meet the clinical cut-off for at least one domain, indicating a significant domain-based impairment; 38/47 (81%) of 16p11.2 deletion, 19/21 (90%) of 16p11.2 duplication, 135/285 (47%) of 22q11.2 deletion and 13/25 (52%) of 22q11.2 duplication carriers. Supplementary Table 4 and Supplementary Figure 1 show for each CNV the proportion of individuals who met the clinical cut-offs for each domain.

#### Autism profiles between genetic variant groups

Genetic variant group predicted autism severity (7% of the variance, p<0.001), autism domain profile (5% of the variance, p<0.001), autism subdomain profile (1% of the variance, p<0.001). In terms of individual domain scores, genetic variant group predicted 5% of the social domain total score, 3% of the communication domain score, and 15% of the RRB domain (Table 2, Figure 1). For subdomain scores the proportion of variance predicted by genetic variant group varied between 1% (social interaction) and 21% (motor mannerisms). In addition to motor mannerisms, the proportion of variance explained was also high for sensorimotor interests (19%). Genetic group variant predicted 12% of variance in FSIQ (p<0.001), 12% of variance in PIQ (p<0.001) and 4% of the variance in VIQ (p<0.001). Findings for autism severity, domain scores and subdomain scores remained significant after controlling for FSIQ, and the eta-squared values remained relatively unchanged (Supplementary Table 5). Age accounted for 0-3% of variance in phenotypic traits (see Supplementary Table 6).

**Table 2:** Comparison of CNV groups on IQ and autism measures. FSIQ, Full Scale Intelligence Quotient; VIQ, Verbal Intelligence Quotient; PIQ, Performance Intelligence Quotient; ADI-R, Autism Diagnostic Interview; RRB, Restricted, Repetitive, and Stereotyped Behaviours. Separate MANCOVA analyses were ran for domain and subdomain scores to avoid including mathematically related scores in the same analysis. Total ADI-R score was analysed separately using an ANCOVA model. Age, gender and site were included as covariates. Post hoc contrasts are given in Supplementary Table 7. **Bold** p-values indicate that the p-value is significant after BH-FDR 0.05 correction *significant after correcting for IQ (Full results in Supplementary Table 3)

|  | 16p11.2 deletion |  |  | 16p11.2 duplication |  |  | 22q11.2 deletion |  |  | 22q11.2 duplication |  |  | Between group variation |  | Within group variation<br>eta-squared % |
| --- | --- | --- | --- | --- | --- | --- | --- | --- | --- | --- | --- | --- | --- | --- | --- |
| Score | N | mean | sd | N | mean | sd | N | score | sd | N | mean | sd | p-value | eta-squared % |  |
| **Cognitive scores** |  |  |  |  |  |  |  |  |  |  |  |  |  |  |  |
| FSIQ | 80 | 81.3 | 15.4 | 50 | 70.9 | 22.4 | 337 | 70.3 | 13.5 | 32 | 88.1 | 21.1 | **<0.001** | 11.7 | 82.2 |
| VIQ | 80 | 78.2 | 18.6 | 49 | 73.6 | 24.4 | 329 | 72.8 | 13.9 | 32 | 84.2 | 18.9 | **<0.001** | 3.7 | 91.7 |
| PIQ | 81 | 85.7 | 15.2 | 50 | 71.9 | 22.4 | 333 | 72.3 | 14.3 | 32 | 87.1 | 21.7 | **<0.001** | 11.8 | 84.8 |
| **ADI-R scores** |  |  |  |  |  |  |  |  |  |  |  |  |  |  |  |
| **Total score** | 82 | 22.0 | 11.8 | 50 | 27.7 | 14.0 | 370 | 16.7 | 11.9 | 45 | 21.2 | 15.4 | **<0.001\*** | 7.4 | 88.7 |
| **Social domain**<br>*subdomains:* | 82 | 12.9 | 7.5 | 50 | 15.8 | 8.0 | 370 | 10.1 | 7.2 | 45 | 12.4 | 9.5 | **<0.001\*** | 5.4 | 91.3 |
| Social interaction | 82 | 2.4 | 1.9 | 50 | 2.9 | 2.0 | 370 | 2.2 | 2.0 | 45 | 2.5 | 2.2 | 0.120 | 1.0 | 96.6 |
| Peer relationships | 82 | 4.2 | 2.3 | 50 | 4.9 | 2.6 | 370 | 3.5 | 2.4 | 45 | 3.8 | 2.8 | **<0.001\*** | 3.1 | 94.2 |
| Shared enjoyment | 82 | 2.5 | 2.2 | 50 | 3.2 | 2.0 | 370 | 1.7 | 1.8 | 45 | 2.5 | 2.4 | **<0.001\*** | 5.7 | 90.1 |
| Socioemotional reciprocity | 82 | 3.9 | 2.5 | 50 | 4.8 | 2.5 | 370 | 2.7 | 2.2 | 45 | 3.7 | 2.9 | **<0.001\*** | 7.9 | 89.8 |
| **Communication domain**<br>*subdomains:* | 82 | 5.8 | 3.7 | 50 | 6.5 | 4.9 | 370 | 4.5 | 3.8 | 45 | 5.4 | 4.3 | **<0.001\*** | 3.1 | 93.6 |
| Gestures | 82 | 2.4 | 2.5 | 50 | 3.1 | 2.9 | 370 | 1.7 | 2.3 | 45 | 2.7 | 2.6 | **<0.001\*** | 3.6 | 94.7 |
| Imagination & Imitation | 82 | 3.5 | 1.8 | 50 | 3.4 | 2.3 | 370 | 2.8 | 2.0 | 45 | 2.7 | 2.1 | **0.009\*** | 2.0 | 92.2 |
| **RRB domain**<br>*subdomains* | 82 | 3.3 | 2.2 | 50 | 5.3 | 2.6 | 370 | 2.1 | 2.3 | 45 | 3.3 | 2.9 | **<0.001\*** | 14.7 | 81.7 |
| Unusual interests | 82 | 0.8 | 1.0 | 50 | 1.6 | 1.1 | 370 | 0.8 | 1.0 | 45 | 0.9 | 1.0 | **<0.001\*** | 4.2 | 92.8 |
| Routines & rituals | 82 | 0.5 | 0.9 | 50 | 1.1 | 1.3 | 370 | 0.5 | 1.0 | 45 | 0.8 | 1.1 | **<0.001\*** | 3.2 | 96.2 |
| Motor mannerisms | 82 | 0.9 | 0.9 | 50 | 1.2 | 0.9 | 370 | 0.2 | 0.5 | 45 | 0.8 | 0.9 | **<0.001\*** | 20.9 | 77.8 |
| Sensorimotor interests | 82 | 1.2 | 0.8 | 50 | 1.5 | 0.7 | 370 | 0.5 | 0.7 | 45 | 0.8 | 0.9 | **<0.001\*** | 18.8 | 74.0 |

**Figure 1:**
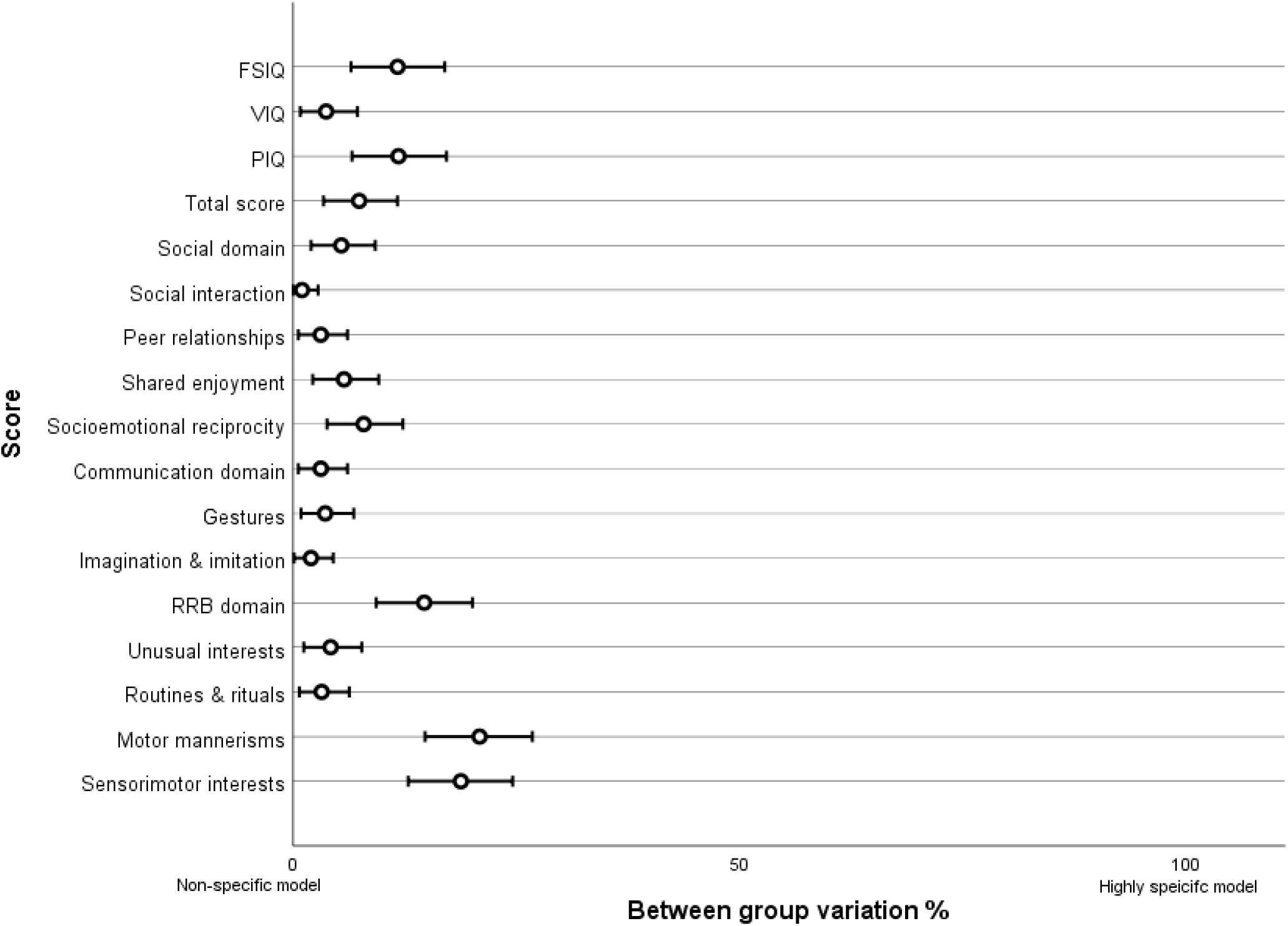
Score variability between genetic variant groups. This plot visualises the between genetic variant group variation data presented in Table 2. Between group eta squared values are plotted on a scale of 0% variance to 100% of the variance. These values represent the proportion of variation in phenotypic outcome predicted by genetic variant group. A value close to 0% would indicate a non-specific model whereby different genotypes lead to similar phenotypic outcomes. As value close to 100% would indicate a highly specific model whereby different genotypes lead to different and discrete phenotypic outcomes. The bars indicate 95% confidence intervals.

After accounting for between group variability, age, gender and site, a large proportion of variability remained; 74% to 97% within group variability depending on trait (final column, Table 2). This is visualised in Figure 1 and Supplementary Figure 2, which shows that although group differences exist, there is much more variability within all groups across traits. For IQ we found greater variability for duplications than deletions for both 16p11.2 (Levene’s test, p=0.001) and 22q11.2 (Levene’s test, p<0.001) loci. For autism severity we found greater variability in outcome for duplications than deletions for the 22q11.2 locus (Levene’s test, p<0.001) but not for the 16p11.2 locus (Levene’s test, p=0.071).

Supplementary Table 7 details which post hoc Tukey contrasts between groups were significant (p-values adjusted for multiple contrasts). To briefly summarise phenotypic profiles; 16p11.2 deletion carriers had relatively moderate autism severity scores and moderate cognitive impairment (IQ= 81.3); 16p11.2 duplication carriers had relatively greater autism severity scores and greater cognitive impairment (IQ= 70.9); 22q11.2 deletion carriers had relatively lower autism severity scores but greater cognitive impairment (IQ=70.3); and 22q11.2 duplication carriers had relatively higher autism severity scores but less cognitive impairment (IQ=88.1).

### Aim 2

Symptom profiles of individuals with autism within the genetic variant groups and individuals with “heterogeneous autism”

Supplementary Table 8 details mean scores for each phenotypic trait for each group (heterogeneous autism, 16p11.2 deletion + autism, 16p11.2 duplication + autism, 22q11.2 deletion + autism, 22q11.2 duplication + autism). With the exception of VIQ and “routines and rituals” domain, all phenotypic traits and subdomains were found to differ between the five groups (last column Supplementary Table 5). These findings remained significant after a B-H FDR 0.05 correction for multiple testing. Age accounted for 0-5% of variance in phenotypic traits (see Supplementary Table 9). Figure 2A visualises the profile of each “genetic variant + autism” group relative to each other and Figure 2B visualises the profile of each “genetic variant+ autism” group relative to the heterogeneous autism group. Supplementary Table 8 details which aspects of the phenotypic profile showed significant contrasts between the heterogeneous autism group and the genetic variant groups.

**Figure 2:**
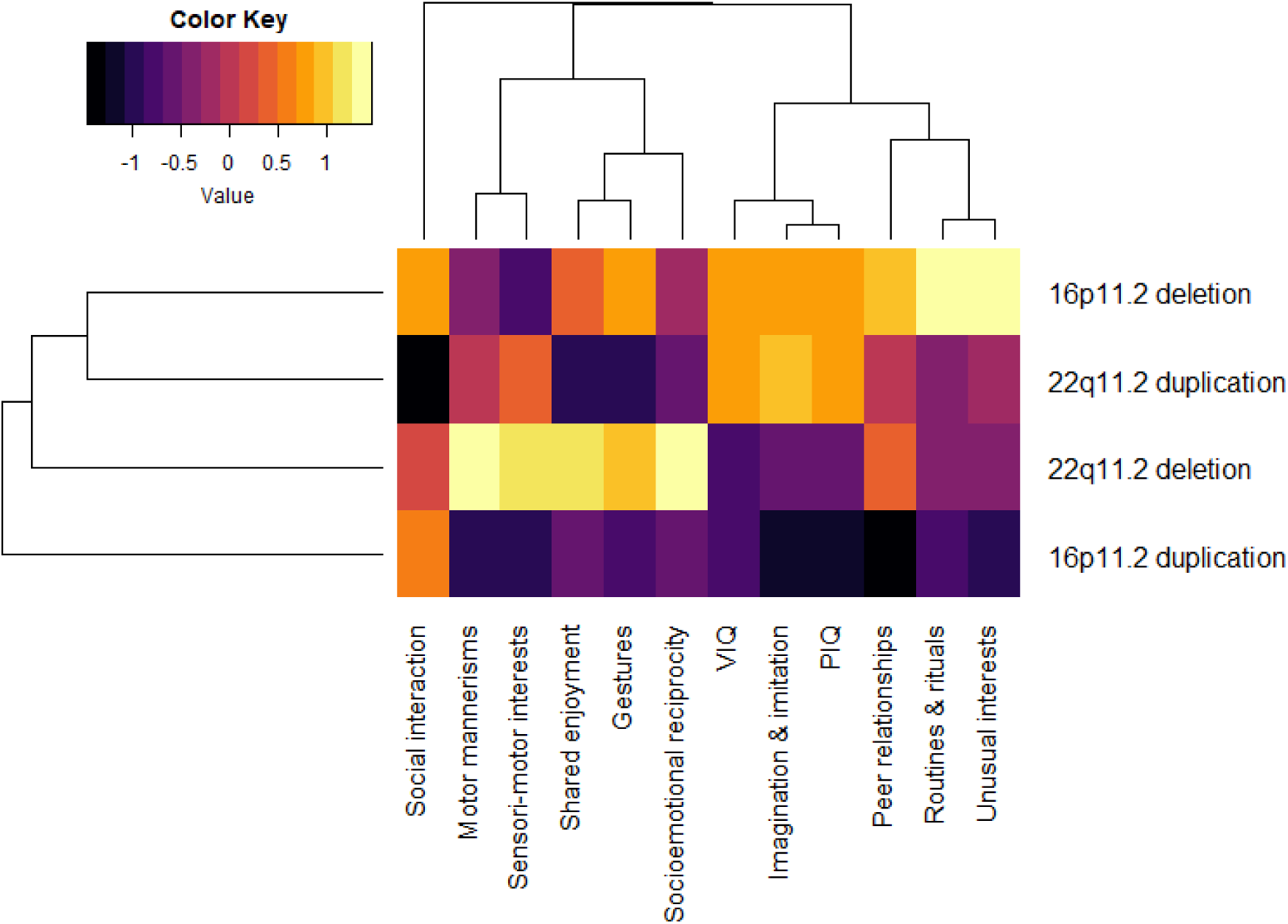
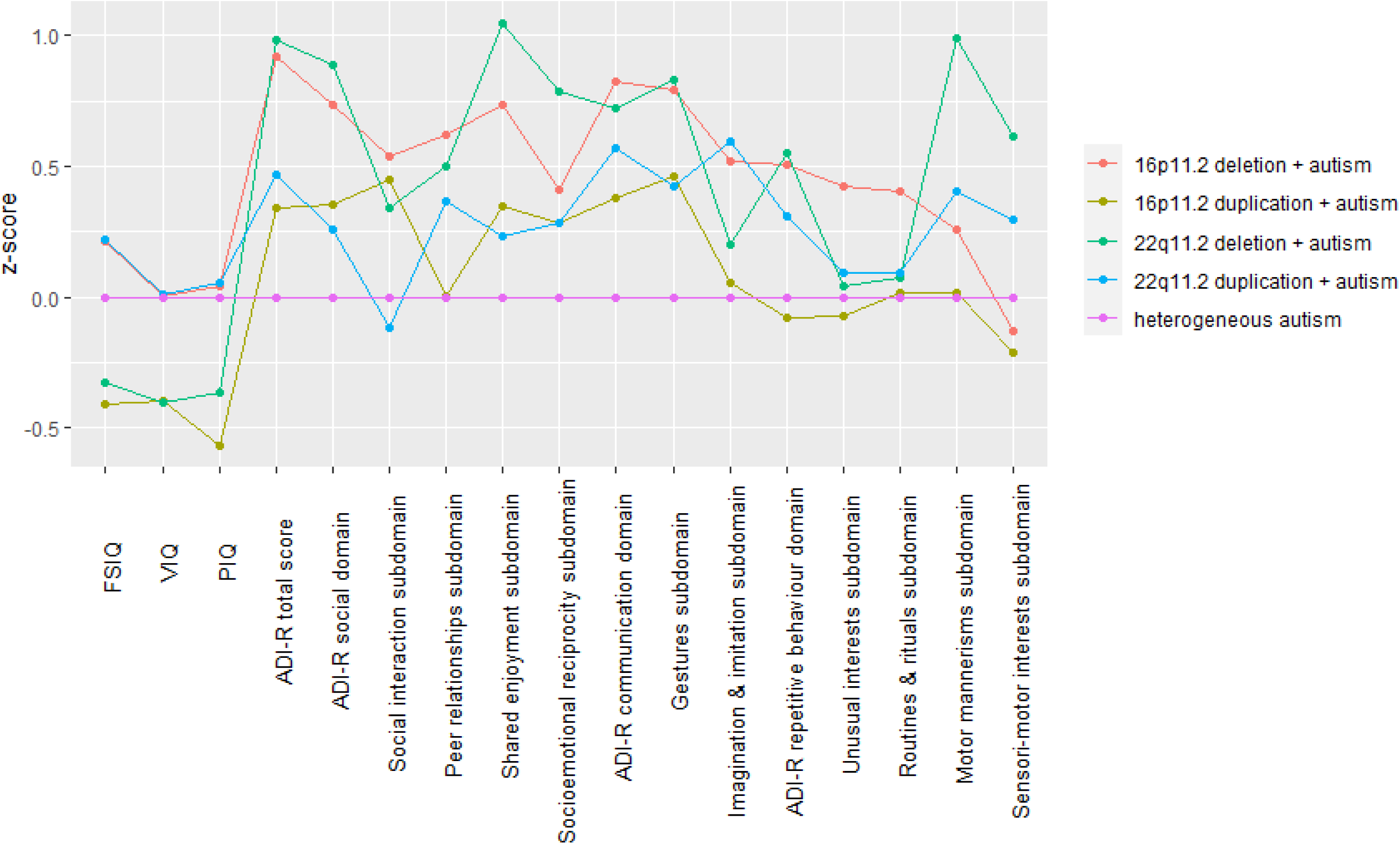
Domain profiles of the genetic variant + autism groups. FSIQ, Full Scale Intelligence Quotient; VIQ, Verbal Intelligence Quotient; PIQ, Performance Intelligence Quotient; ADI-R, Autism Diagnostic Interview. **2A** To visualise how the “genetic variant + autism” groups differed, a heatmap plot was generated by transforming IQ and ADI-R scores of the “genetic variant + autism” groups to z-scores, dendrograms showing the clustering of CNVs and phenotypes were generated using methods described for 3A. Scores for each “genetic variant + autism” were standardized into z scores relative to each other and were adjusted for sex, age and site. The z scores were constructed so that a negative score always denoted a poorer performance. Black indicates a relative deficit in that neuropsychiatric domain compared to other CNV carriers, yellow represents a relative strength compared to other CNV carriers. Hierarchical clustering, for the purposes of presentation (indicated by the dendrogram), was performed using Ward’s method and Euclidian distance. **2B** To visualise the profiles of the “genetic variant + autism” groups relative to the heterogeneous autism group phenotypic scores were standardised to z-scores, using the mean and SD of the heterogeneous autism group as reference—i.e., the difference in the individual’s score and the mean score for the entire autism heterogeneous group was divided by the SD for the heterogeneous autism group. The z-scores were adjusted for sex, age and site. We constructed these Z scores so that a negative score for a -CNV carrier indicated a worse outcome.

To briefly summarise phenotypic profile differences relative to the heterogeneous autism group: the 16p11.2 deletion + autism group had relatively less impairment in autism score severity but had a similar level of cognitive impairment; the 16p11.2 duplication + autism group had greater PIQ deficits but did not differ on any of the other phenotypic measures; the 22q11.2 deletion + autism group had greater cognitive impairment but relatively less severity in autism scores; the 22q11.2 duplication + autism group did not significantly differ from the heterogeneous autism group on any phenotypic measure.

#### Sex

Male CNV carriers (all groups combined) were at increased risk of autism (OR = 2.3, p<0.001) compared to female CNV carriers. However, male to female ratios were lower within CNV carriers with autism (2.3:1) compared to the heterogeneous autism group (6.4:1) (p<0.001).

## Discussion

This study is the result of a collaboration between several international genetics-first consortia and the Autism Genome Project. The availability of a large sample of individuals with one of four autism risk CNVs allowed us to use a genetics-first approach which meant we were not constrained by ascertaining patients on the basis of autism diagnosis, allowing examination of the impact of genotype on autism severity and domain profiles across the spectrum. The use of the widely accepted research diagnostic ADI-R interview across all sites represents a methodological strength, enabling us to directly compare the autism profiles of 22q11.2 and 16p11.2 CNVs. Our findings indicate that although autism risk genetic variants differ in several aspects of the autism phenotype, including autism severity, symptom domain profile and cognitive profile, only 1-21% of the variance is explained by genetic variant group, depending on autism measure. In contrast, variation within each of the four genetic variant groups is much greater, explaining between 74%-97% of the variability, depending on autism measure. This highlights that even within individuals with the same autism risk genetic variant, the autism profile is difficult to predict on the basis of CNV alone and that phenotypic profiles overlap, providing evidence against a ‘highly specific” model ^42^ whereby each genotype leads to a unique autism phenotype (see Supplementary Figure 3), instead our findings support a partially specific model whereby autism profiles are distinct but overlapping.

Severity of autism phenotype differed by genetic variant group. In terms of autism prevalence; fewer 22q11.2 deletion carriers met criteria for autism (23%) than 22q11.2 duplication (44%), 16p11.2 deletion (43%) and 16p11.2 duplication (58%) carriers. These figures represent autism prevalence within a clinically ascertained cohort of CNV carriers, and should not be taken as the prevalence for CNV carriers in the wider population. Among CNV carriers with autism we found that 22q11.2 deletion and 16p11.2 deletion carriers with autism had relatively less severe profiles compared to individuals with heterogeneous autism. On the other hand individuals with 16p11.2 duplication and 22q11.2 duplication with autism had a profile more consistent with individuals with heterogeneous autism. Our findings complement genome wide CNV studies which find the strength of association and penetrance for autism varies by genetic variant, in particular the association of 22q11.2 deletion is relatively weaker^8^.

We found evidence that the four genetic variant groups were associated with differences in autism severity, the three autism domains as well as nine out of the 10 subdomains we studied, FSIQ, VIQ and PIQ. However, the proportion of variance explained by genetic variant group for each sub-domain varied between 1-21%. It was only the social interaction subdomain that did not differ, indicating that this trait is a universal aspect of autism across the 4 genetic variant groups. The sub-domains for which genetic variant group explained the greatest proportion of variance were motor aspects of the RRB domain, motor mannerisms (21%) and sensorimotor interests (19%), indicating that genetic variant group particularly distinguishes motor aspects of the autism phenotype. Cognitive profile was also influenced by genetic variant group; 22q11.2 deletion and 16p11.2 duplication carriers had greater cognitive impairments in FSIQ, VIQ and PIQ relative to 22q11.2 duplication and 16p11.2 deletion carriers. There was evidence at both the 22q11.2 and 16p11.2 loci that cognitive outcomes are more variable for duplication carriers than deletion carriers. This has been previously been reported for 16p11.2 duplication carriers^25^ and our findings indicate the same may be true for the 22q11.2 locus. Autism severity of a genetic variant did not covary with magnitude of cognitive deficit, 22q11.2 duplication carriers had the highest mean IQ (88.1) out of the CNV groups, yet had high symptom severity scores. 22q11.2 deletion carriers had the greatest cognitive impairment yet were at less risk of autism compared to the other genetic variants. Furthermore, when we controlled for IQ, differences in autism domain and subdomains scores between CNVs remained relatively unchanged. These findings suggest that the mechanisms underlying autism and cognitive impairment are at least partially distinct among carriers of pathogenic CNVs.

However, although specific group differences exist, it is clear that phenotypic profiles overlap (Supplementary Figure 2), and we find greater variability between individuals with the same CNV than between CNVs. Overall, our findings provide most support for a “partially specific model” whereby autism profiles are distinct but highly overlapping. Though the magnitude of these differences is closer to the “non-specific effect” end of the scale whereby all genotypes lead to similar autism phenotypes, than the “highly specific effect” end of the scale whereby genotypes lead to discrete autism subtypes (Figure 1). These findings highlight that it will be important for behavioural phenotyping research to move beyond a focus on average differences between variants, and to investigate the genetic (including additional rare variants and polygenic risk, which we were not able to analyse in this study) and environmental factors that contribute to variation in clinical phenotypes. There is already evidence that family background is important to consider in a genetic counselling context; parental IQ has been found to predict the IQ impairment in 16p11.2 and 22q11.2 deletion carriers^43-45^.

There was a male preponderance for autism across all genetic variant groups, and gender significantly influenced domain and subdomain profiles. However, the male to female ratio in CNV carriers is approximately 2.3:1 which is considerably less pronounced than in the heterogeneous autism group (6.4:1). It may be that the genetic variants we studied have such a large effect on neurodevelopment that they partially override the protective effect of being female^9,11^. Age did influence phenotypic traits, however the proportion of variance age explained in analyses was low (≤5%).

Using a genetics-first approach, we identified a significant proportion of CNV carriers (54%) who did not meet autism criteria but did meet clinical cut-offs for diagnosis related impairments. Furthermore, the profile of CNV carriers with autism does to some extent present differently from heterogeneous autism (Figure 2B). This has the potential implication that the clinical needs of patients with genomic conditions may be overlooked because they fail to meet diagnostic criteria despite exhibiting a range of impairments across domains. Parents of children with CNVs at 16p11.2 or 22q11.2 who have taken part in our studies in the UK have anecdotally reported that their child’s genetic diagnosis can be a barrier to receiving an autism diagnosis and support, with some service providers having stated that a child with a genetic diagnosis cannot also have a secondary diagnosis of autism despite DSM 5 specifying that autism can be diagnosed when “associated with a known medical or genetic condition or environmental factor”^1^. It is important that clinicians are aware of the risk of autism associated with certain genetic variants to improve the opportunities that these children receive of an early diagnosis and access to interventions.

Further clinical implications arise from our finding that there are not highly specific genotype-phenotype relationships between individual CNVs and autism, at least for 16p11.2 and 22q11.2 deletion and duplication variants. This indicates that, although CNVs are pre-symptomatically predictive of autism and therefore can inform early intervention, individual genotypes are not specific in predicting symptom subtypes. Rather our findings indicate an overlap in clinical phenotypes between these CNVs, suggesting that neurodevelopmental service provision for different CNVs could be grouped together. Our genetics-first approach reveals great variability within CNV groups, highlighting that autism risk variants are not deterministic for autism. It is important that in genomic counselling that pathogenic CNVs are considered as one factor within a broader biopsychosocial context, rather than being the only causative factor for autism. Identification of genetic and environmental modifiers of phenotypes of autism risk CNVs has potential for informing clinical care and intervention.

Our study benefits from several features, including a large sample size by combining data from individuals with these rare genetic conditions from a number of international cohorts, and synchronisation of phenotyping measures across sites allowing for analysis extending beyond categorical diagnosis, allowing for autism domains and subdomains to be analysed. However, there are potential limitations. Firstly, ascertainment bias needs to be considered as our study focuses on individuals who received a clinical genetic diagnosis, and our findings therefore do not necessarily extend to individuals with these CNVs in the population who are affected below a clinical threshold and as a consequence not referred for genetic testing. As one of the main indications for genetic testing currently is often developmental delay^24,25^, our findings may not be representative for individuals with these CNVs with a more typical developmental pattern. However, despite these ascertainment considerations, not all CNV carriers in this study met autism criteria or had cognitive impairment, thus allowing us to study the impact of genotype across a broad spectrum of abilities. Another source of possible ascertainment bias is that referral reason for genetic testing may differ by genetic variant. For instance it has been reported that 22q11.2 deletion carriers are more likely to be referred due to physical abnormalities, such as heart defects, in comparison to 22q11.2 duplication carriers who are more likely to be referred for developmental reasons^24^. However, this may actually reflect true phenotypic differences as a recent population based study which was able to identify individuals in the population undiagnosed with a 22q11.2 CNV, as well as individuals with a diagnosis through a clinic, reported higher frequency of congenital abnormalities in the deletion carriers^46^. Before taking part in the study, individuals had a variety of diagnostic experiences, where some had a pre-existing autism diagnosis before the ADI-R assessment, whilst others had had no interaction with autism diagnostic services. This potentially introduces caregiver reporter bias, but this is partly mitigated by the semi-structured nature of the ADI-R. That is, although the ADI-R interview is based on caregiver report, the scoring of a particular trait is based on concrete descriptions coded by a trained interviewer. We were not able to conduct cross-site reliability of ADI-R administration as it was not pre-planned that ADI-R data would be combined across several international sites, however all assessors underwent ADI-R formal training and were research reliable. Finally, we were not able to control for ethnicity, and socio-economic and environmental factors, as these data were not available at all sites, and/or were not internationally comparable. Future studies would benefit from greater alignment of measurement of environmental factors across international sites.

## Conclusion

The genetics-first approach we employed represents a novel method for investigating genotype-phenotype relationships unconstrained by categorical diagnostic criteria. We found that the phenotypic profiles of 16p11.2 and 22q11.2 CNVs differ in terms of severity, symptom profile and cognitive profile. However, although genetic variants have specific effects, within variant variability is much greater than between variant variability, thus indicating that the phenotypic consequences of genomic risk factors for autism fit a “partial specific model” rather than a “highly specific” model. It will be important that future studies of autism risk variants consider the genetic and environmental factors that contribute to clinical variability within autism risk variant carriers. An important message from our work is that individuals with genomic conditions are likely to present with clinically significant symptoms of autism but not meet diagnostic criteria. Clinical services need to adapt as individuals without a formal autism diagnosis are unlikely to access support and interventions.

## Data Availability

Due to ethical restrictions, data is not available to be shared.

## Acknowledgements

We thank all the children and families who took part in this study, the UK National Health Service (NHS) medical genetic clinics for their support, and charities including Unique, Max Appeal and 22Crew for their support. We thank all members of the IMAGINE-ID consortium for their contributions. We thank the core laboratory team of the Division of Psychological Medicine and Clinical Neurosciences laboratory at Cardiff University (Cardiff, UK) for DNA sample management and genotyping. We are grateful to all of the families at the participating Simons VIP sites, as well as the Simons VIP Consortium. We appreciate obtaining access to the phenotypic data on SFARI Base. Approved researchers can obtain the Simons VIP (now known as Simons Searchlight) population dataset described in this study by applying at https://base.sfari.org. We also thank the International 22q11.2 Brain and Behaviour Consortium and the National Centre for Mental Health, a collaboration between Cardiff, Swansea and Bangor Universities, for their support.

