## Supplementary Materials for "A Genetics-First Approach to Dissecting the Heterogeneity of Autism: Phenotypic Comparison of Autism Risk Copy Number Variants"

**Supplementary Table 1:** Number of participants at each site

|  |  | **Number of individuals contributed** | | | |
| --- | --- | --- | --- | --- | --- |
|  | **Research Centre** | **16p11.2 deletion** | **16p11.2 duplication** | **22q11.2 deletion** | **22q11.2 duplication** |
|  | Cardiff University ECHO & IMAGINE-ID cohorts | 8 | 8 | 67 | 11 |
|  | Belgrade University Children’s Hospital |  |  | 8 |  |
|  | Children’s Hospital of Philadelphia |  |  |  | 18 |
|  | 16p11.2 European consortium | 4 | 9 |  |  |
|  | Simons VIP | 70 | 33 |  |  |
|  | University of California, Los Angeles |  |  | 69 | 16 |
|  | University Medical Center Utrecht |  |  | 140 |  |
|  | The State University of New York |  |  | 86 |  |

**Supplementary Table 2:** Genotype information

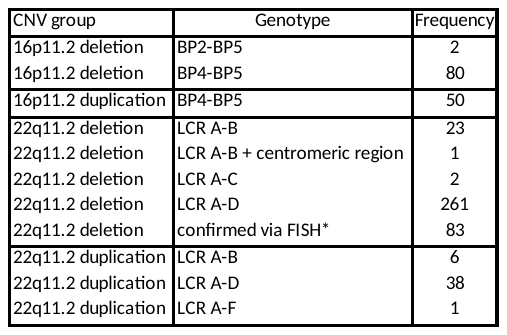

BP, breakpoints; LCR, low copy repeat region; FISH, Fluorescence in situ hybridization.

*22q11.2 deletion confirmed by FISH, indicating deletion hits the critical region but exact LCRs could not be defined.

**ADI-R**

Following previous studies^27,34,45^ we only analysed the scores that contributed to the diagnostic algorithm, the majority of which are early developmental “ever/most abnormal” items cohort^34^. The social domain comprises four domains: ‘social interaction’, ‘peer relationships’, ‘shared enjoyment’ and ‘socioemotional reciprocity’ subdomains. For the communication domain we excluded subdomains that were designed only for verbal individuals and only retained those available for both verbal and nonverbal individuals, which were ‘gesture’ and ‘imitation and imagination’ subdomains. This followed the approach of a previous study that examined subdomain scores^45^. The RRB domain comprises ‘repetitive behaviour’, ‘routines and rituals’, ‘motor mannerisms’ (hand and finger mannerisms, and stereotyped body movements) and ‘sensorimotor interest’ (unusual sensory interests and repetitive use of objects) subdomains. All ADI-R assessors were extensively trained to research reliability standards and each site used consensus coding procedures.

**Supplementary Table 3**: ADI-R assessors and sites of assessment

| Site | Assessors | Sites of assessments |
| --- | --- | --- |
| Cardiff University | Dr Samuel Chawner (postdoctoral research psychologist), Hayley Moss (psychology research assistant), Dr Joanne Doherty (Child and Adolescent Mental Health Services psychiatrist). Training and reliability by certified ADI-R trainer Dr Sarah Curran (Consultant Child and Adolescent Mental Health Services Psychiatrist). | Assessments administered in the patients home, or at the Hadyn Ellis Building clinic at Cardiff University. |
| Belgrade University Children’s Hospital | Milica Pejovic-Milovancevic (Child psychiatrist, Institute of Mental Health, Belgrade)  Marina Mihaljevic (Psychiatrist, Clinic for Psychiatry, Clinical Center of Serbia). Assessors were trained by ADI-R trainer Prof Patrick Bolton Consultant Child and Adolescent Mental Health Services Psychiatrist | Assessments administered at the Institute of Mental Health, Belgrade. |
| Children’s Hospital of Philadelphia | Research-reliable, Licensed Ph.D. or supervised M.A. clinical psychologists. | Assessments administered remotely from Children’s Hospital of Philadelphia |
| 16p11.2 European consortium | Trained and certified psychologists. ADI-R training provided by Professor Bernadette Rogé. | Assessments administered at Lausanne University hospital |
| Simons VIP | Research-reliable, Licensed Ph.D. or supervised M.A. clinical psychologists. Training and reliability were conducted by Catherine Lord’s lab (UMACC/CADB) and/or independent certified ADI-R trainers. | Assessments administered at one of five Simons VIP sites  (Baylor College of Medicine, Boston Children’s Hospital, Children’s  Hospital of Philadelphia, the University of California San Francisco, and the University of Washington). |
| University of California, Los Angeles | Clinical psychologists and psychometrists trained I n the ADI-R to research reliability levels by the UCLA Center for Autism Research and Treatment (CART). | Assessments administered at University of California Los Angeles, Semel Institute for Neuroscience and Human Behavior. |
| University Medical Center Utrecht | M.D. PhD child/adolescent psychiatrist and M.Sc. psychologist, who were certified in ADI-R assessments. Training and reliability were conducted  by an independent,   certified ADI-R trainer. | Assessments administered took place at the psychiatry department of the University Medical Center Utrecht. |
| The State University of New York - Upstate Medical University | Research-reliable, licensed Ph.D. clinical child psychologist and M.D. child/adolescent psychiatrist. | Assessments administered at State University of New York, Upstate Department of Psychiatry. |

**Analysis of covariance models**

*Autism severity:* ANCOVA with ADI-R total score as the phenotypic outcome. *Autism domain profile:* multivariate analysis of covariance (MANCOVA) with social, communication and repetitive behaviour ADI-R domain total scores as the phenotypic outcomes entered into the same model. *Autism subdomain profile:* MANCOVA model with ADI-R social subdomains (social interaction, peer relationships, shared enjoyment and socioemotional reciprocity), ADI-R communication subdomains (gestures, imagination and imitation), ADI-R repetitive behaviour subdomains (unusual interests, routines and rituals, motor mannerisms and sensorimotor mannerisms) as phenotypic outcomes entered into the same model. *Cognitive profile:* ANCOVA model with FSIQ as the phenotypic outcome, and a further MANCOVA model with PIQ and VIQ as phenotypic outcomes entered into the same model.

**Supplementary Table 4: Proportion of CNV carriers who met clinical cut-offs for the three autism domains**

|  | Social | Communication | RRB |
| --- | --- | --- | --- |
| 16p11.2 deletion | 60% | 73% | 59% |
| 16p11.2 duplication | 72% | 74% | 86% |
| 22q11.2 deletion | 47% | 46% | 32% |
| 22q11.2 duplication | 53% | 64% | 58% |

RRB: repetitive and restrictive behaviour

**Supplementary Figure 1: Proportion of CNV carriers who met clinical cut-offs for the three autism domains**

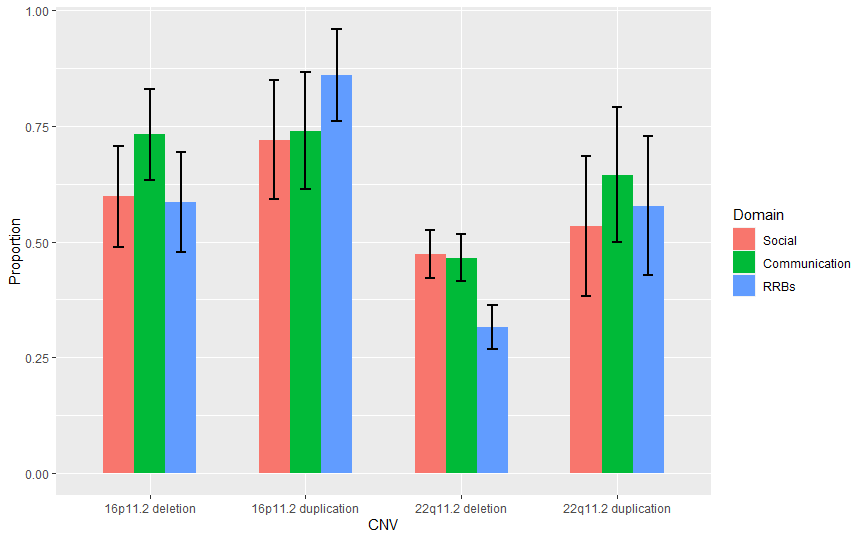

Error bars indicate 95% confidence intervals. RRB: repetitive and restrictive behaviour.

**Supplementary Figure 2: Distribution of total ADI-R score, autism domain scores, and FSIQ by genetic variant group**

The range of scores for total ADI-R scores is highly overlapping between groups (16p11.2 deletion, 3 to 48; 16p11.2 duplication, 0 to 53; 22q11.2 deletion, 0 to 48; 22q11.2 duplication, 0 to 48). Similarly, the IQ ranges overlap (16p11.2 deletion, 42 - 122; 16p11.2 duplication, 28 - 108; 22q11.2 deletion, 42 - 117; 22q11.2 duplication, 30 - 119).

For IQ we found greater variability for duplications than deletions for both 16p11.2 (Levene’s test, p=0.001) and 22q11.2 (Levene’s test, p<0.001) loci. For autism severity we found greater variability in outcome for duplications than deletions for the 22q11.2 locus (Levene’s test, p<0.001) but not for the 16p11.2 locus (Levene’s test, p=0.071).

Straight lines indicate group mean.
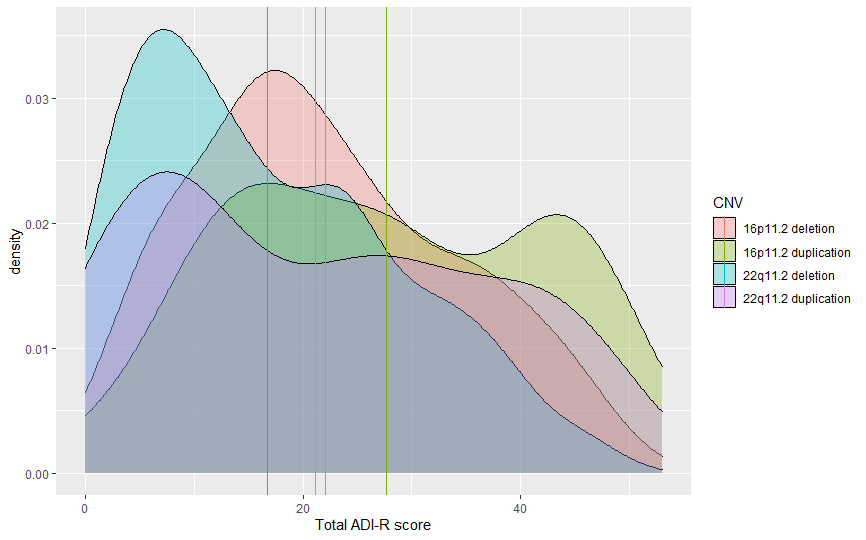

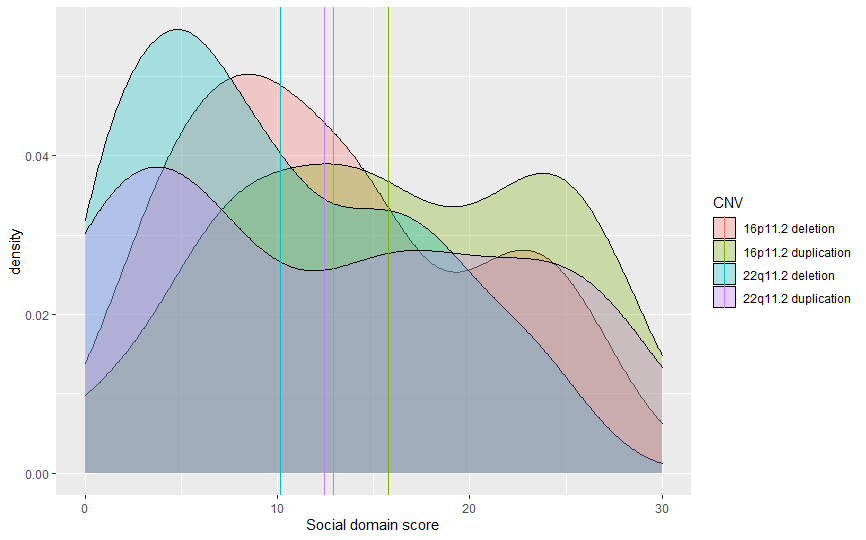

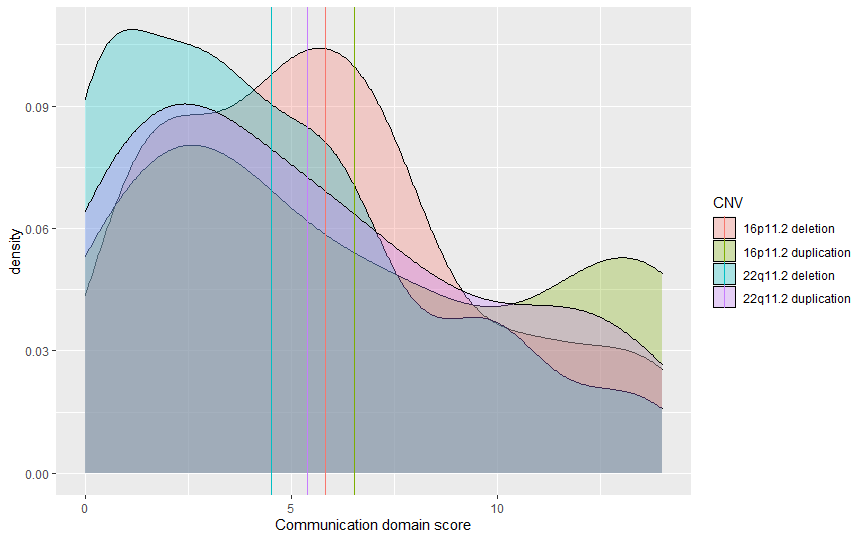

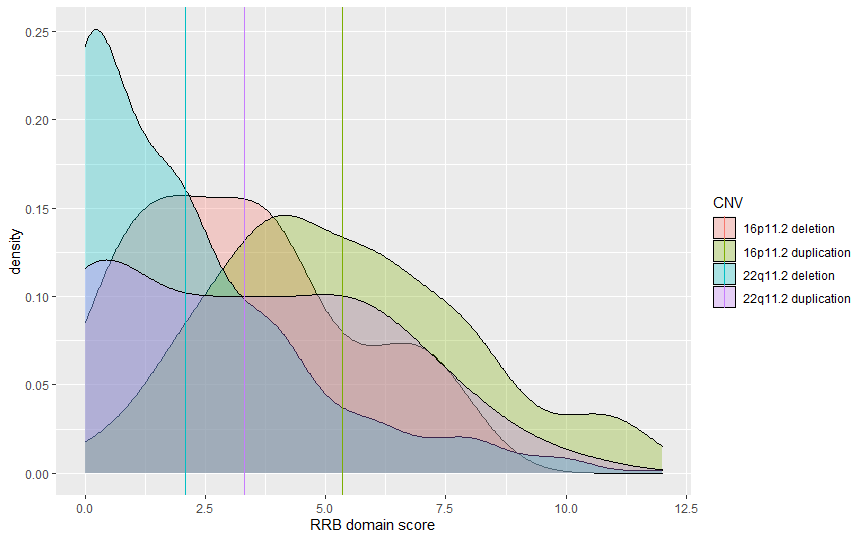

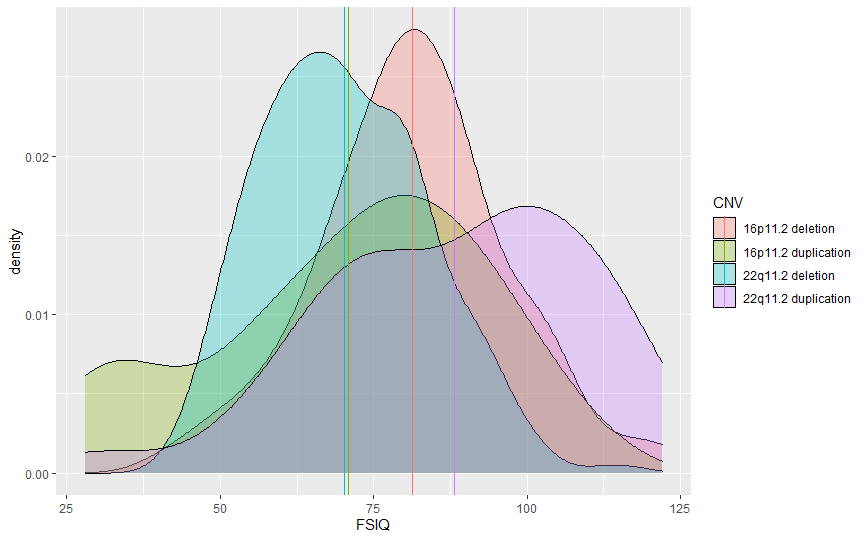

**Supplementary Table 5:** Between group variance adjusted for FSIQ

FSIQ, Full Scale Intelligence Quotient; VIQ, Verbal Intelligence Quotient; PIQ, Performance Intelligence Quotient ; ADI-R, Autism Diagnostic Interview ; RRB, Restricted, Repetitive, and Stereotyped Behaviours.

|  | Score | Between group variation controlled for FSIQ | |
| --- | --- | --- | --- |
|  | **ADI-R scores** | p-value | eta-squared % |
|  | **Total score** | <0.001 | 8.1 |
|  | **Social domain** | <0.001 | 6.0 |
|  | *subdomains:* |  |  |
|  | Social interaction | 0.044 | 1.6 |
|  | Peer relationships | <0.001 | 3.3 |
|  | Shared enjoyment | <0.001 | 5.7 |
|  | Socioemotional reciprocity | <0.001 | 8.5 |
|  | **Communication domain** | <0.001 | 3.2 |
|  | *subdomains:* |  |  |
|  | Gestures | <0.001 | 3.5 |
|  | Imagination & Imitation | 0.011 | 2.0 |
|  | **RRB domain** | <0.001 | 16.4 |
|  | *subdomains* |  |  |
|  | Unusual interests | <0.001 | 4.9 |
|  | Routines & rituals | <0.001 | 3.5 |
|  | Motor mannerisms | <0.001 | 23.4 |
|  | Sensorimotor interests | <0.001 | 19.9 |

**Supplementary Table 6:** Effect of age in Aim 1 MANCOVA analyses

FSIQ, Full Scale Intelligence Quotient; VIQ, Verbal Intelligence Quotient; PIQ, Performance Intelligence Quotient ; ADI-R, Autism Diagnostic Interview ; RRB, Restricted, Repetitive, and Stereotyped Behaviours.

|  | Score | p-value | Eta-squared | Fixed effect slope coefficient |
| --- | --- | --- | --- | --- |
|  | **Cognitive scores** |  |  |  |
|  | FSIQ | 0.009 | 1.2 | -0.37 |
|  | VIQ | 0.088 | 0.6 | -0.27 |
|  | PIQ | <0.001 | 1.9 | -0.54 |
|  | **ADI-R scores** |  |  |  |
|  | **Total score** | 0.232 | 0.2 | -0.18 |
|  | **Social domain** | 0.233 | 0.2 | -0.11 |
|  | *subdomains:* |  |  |  |
|  | Social interaction | 0.921 | 0.0 | -0.01 |
|  | Peer relationships | 0.106 | 0.5 | -0.05 |
|  | Shared enjoyment | 0.570 | 0.1 | 0.01 |
|  | Socioemotional reciprocity | 0.014 | 1.0 | -0.07 |
|  | **Communication domain** | 0.766 | 0.0 | -0.02 |
|  | *subdomains:* |  |  |  |
|  | Gestures | 0.215 | 0.3 | -0.03 |
|  | Imagination & Imitation | 0.348 | 0.2 | 0.01 |
|  | **RRB domain** | 0.055 | 0.6 | -0.05 |
|  | *subdomains* |  |  |  |
|  | Unusual interests | 0.315 | 0.2 | 0.01 |
|  | Routines & rituals | 0.197 | 0.3 | -0.01 |
|  | Motor mannerisms | 0.132 | 0.3 | -0.01 |
|  | Sensorimotor interests | <0.001 | 2.9 | -0.03 |

**Supplementary Table 7:** Phenotypic posthoc contrasts between genetic variant groups

Arrows indicate the direction of effect of the difference between genetic variant groups; an upward arrow ↑ indicates that the first group mentioned in the pair has a higher value than the second group mentioned; a downward arrow ↓ indicates the opposite; no arrow indicates that the values were the same between groups. For example for FSIQ, 16p11.2 duplication carriers have a lower FSIQ than 16p11.2 deletion carriers. Tukey’s method was used to conduct posthoc contrasts between CNV groups, producing p-values adjusted for the number of contrasts. Green highlighted cells indicate a significant contrast.

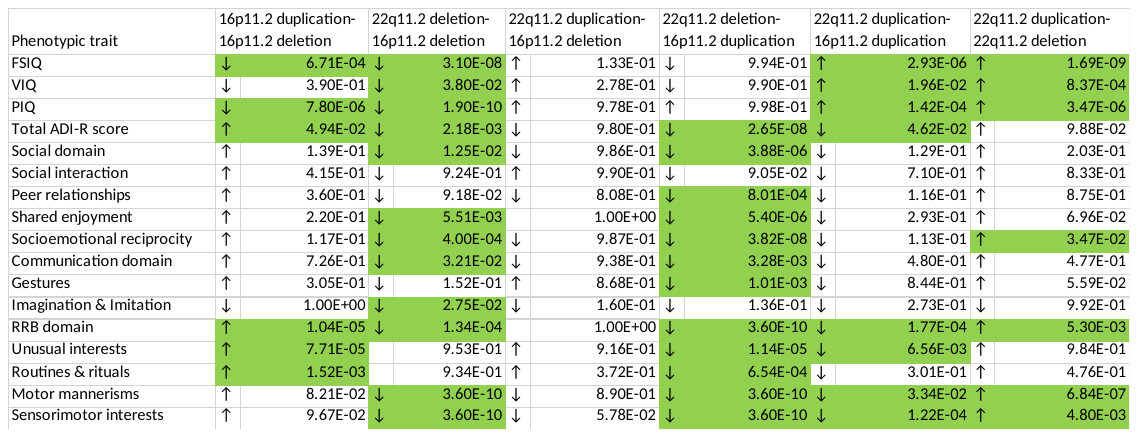

**Supplementary Table 8:** Comparison of autism in the genetic variant groups to the heterogeneous autism group in terms of IQ and symptom domains

FSIQ, Full Scale Intelligence Quotient; VIQ, Verbal Intelligence Quotient; PIQ, Performance Intelligence Quotient ; ADI-R, Autism Diagnostic Interview.

Separate MANCOVA analyses were ran for domain and subdomain scores to avoid including mathematically related scores in the same analysis. Total ADI-R score was analysed separately using an ANCOVA model. Age, gender and site were included as covariates.

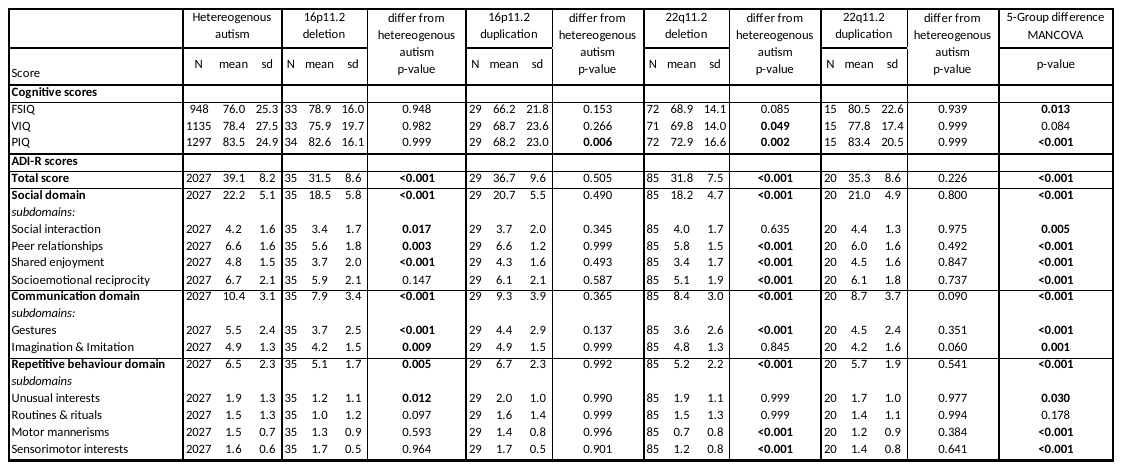

**Supplementary Table 9:** Effect of age in Aim 2 MANCOVA analyses

FSIQ, Full Scale Intelligence Quotient; VIQ, Verbal Intelligence Quotient; PIQ, Performance Intelligence Quotient ; ADI-R, Autism Diagnostic Interview ; RRB, Restricted, Repetitive, and Stereotyped Behaviours.

|  | Score | p-value | Eta-squared | Fixed effect slope coefficient |
| --- | --- | --- | --- | --- |
|  | **Cognitive scores** |  |  |  |
|  | FSIQ | **<0.001** | 1.8 | 0.31 |
|  | VIQ | **<0.001** | 3.6 | 0.88 |
|  | PIQ | 0.568 | 0.0 | -0.22 |
|  | **ADI-R scores** |  |  |  |
|  | **Total score** | **<0.001** | 2.1 | 0.24 |
|  | **Social domain** | **<0.001** | 1.9 | 0.14 |
|  | *subdomains:* |  |  |  |
|  | Social interaction | **<0.001** | 5.3 | 0.07 |
|  | Peer relationships | **0.007** | 0.3 | 0.02 |
|  | Shared enjoyment | **<0.001** | 1.5 | 0.04 |
|  | Socioemotional reciprocity | 0.114 | 0.1 | 0.01 |
|  | **Communication domain** | **<0.001** | 1.0 | 0.06 |
|  | *subdomains:* |  |  |  |
|  | Gestures | **0.008** | 0.3 | 0.03 |
|  | Imagination & Imitation | **<0.001** | 1.7 | 0.04 |
|  | **RRB domain** | **<0.001** | 0.6 | 0.03 |
|  | *subdomains* |  |  |  |
|  | Unusual interests | **<0.001** | 1.7 | 0.03 |
|  | Routines & rituals | **<0.001** | 1.0 | 0.03 |
|  | Motor mannerisms | **<0.001** | 0.8 | -0.01 |
|  | Sensorimotor interests | **<0.001** | 0.6 | -0.01 |

**Supplementary Figure 3:** Hypothetical plots of “non-specific”, “partially specific” and “highly specific” models of genotype-phenotype relationships within autism

We conceptualise three possible models (Figure 1); the “nonspecific effect” model whereby all genotypes lead to similar autism phenotypes; the “highly specific” model whereby each genotype leads to a unique autism phenotype; and the “partially specific model” whereby autism profiles are distinct but overlapping.

Non-specific model: for Trait A, genotype does not explain any of the variance in phenotype, indicating that phenotypic variability is explained by within group factors. Highly specific model: for Trait C genotype explains all the variability in phenotypic outcome, indicating that there is no within group variability. Partially specific model: where both between genotype and within genotype variability exist.

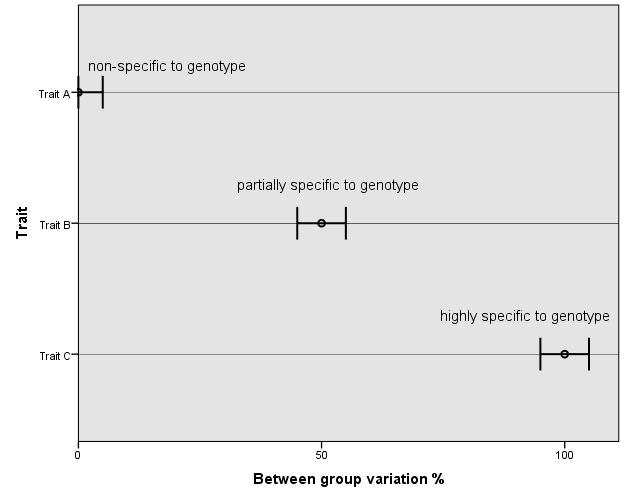
